# Addressing the aftermath of the COVID-19 pandemic: A quality improvement collaborative to optimize the use of antibacterials in Argentine Intensive Care Units

**DOI:** 10.1101/2023.12.28.23300542

**Authors:** Facundo Jorro-Barón, Cecilia Loudet, Wanda Cornistein, Inés Suárez-Anzorena, Pilar Arias-López, Carina Balasini, Laura Cabana, Eleonora Cunto, Rodrigo Corral, Luz Gibbons, Marina Guglielmino, Gabriela Izzo, Marianela Lescano, Claudia Meregalli, Cristina Orlandi, Fernando Perre, María Elena Ratto, Mariano Rivet, Ana Paula Rodríguez, Viviana M. Rodríguez, Paula Romina Villegas, Emilse Vitar, Javier Roberti, Ezequiel García-Elorrio, Viviana E. Rodriguez

## Abstract

**Background:** Reducing antimicrobial resistance is a global priority that become even more important after the COVID-19 pandemic. To date there is a scarce volume of evidence from antimicrobial stewardship programs from less resourced settings where this phenomenon is bigger. Our aim was to improve the quality of antibacterials prescription in intensive care units (ICUs) in a middle-income country.

**Methods:** We established a quality improvement collaborative (QIC) model involving nine ICUs over an 11-month period, with a 16-week baseline (BP) and 32-week Intervention (IP) periods. Our co-designed intervention package included audits and feedback on antibacterial use, facility-specific treatment guidelines, antibacterial timeouts, pharmacy-based interventions, and education. The intervention was delivered in two learning sessions with three action periods, along with coaching support and basic quality improvement training.

**Results:** We enrolled 912 patients, with 357 in baseline period (BP) and 555 in implementation period (IP). The latter had higher APACHE II (17 (12, 21) vs. 15 (11, 20); p=0.036) and SOFA scores (6 (4, 9) vs. 5 (3, 8); p=0.006), sepsis (36.1% vs. 31.6%, p<0.001), and septic shock (40.0% vs. 33.8%, p<0.001). Days of antibacterial therapy were similar between groups (IP 1112.2, BP 1133.4, RR 0.98 (0.95-1.02); p=0.2973) and the antibacterial Daily Define Dose was lower in IP group (IP, 1193.0; BP, 1301.0; RR, 0.92 (0.89, 0.95); p=0.0001). The rate of adequate antibacterial adjustment was higher during the IP (62.0% vs. 45.3%, p<0.001). We observed a lower rate of ventilation-associated pneumonia and catheter-associated urinary tract infections related to multidrug-resistant organisms (MDRO) in the IP. There was a noticeable improvement in the Infection Prevention and Control (IPC) Assessment Framework compared to baseline.

**Conclusion:** The implementation of a post pandemic antimicrobial stewardship program in ICUs via a QIC demonstrated success in improving antibacterials utilization, reducing HAIs related to MDRO while also enhancing IPC measures.

**What is already known on this topic:**

- Healthcare-associated infections represent a global healthcare issue, particularly prevalent in low- and middle-income countries, where their occurrence is nearly three times higher.
- Approximately 50% of antimicrobial use is deemed unnecessary or inappropriate, necessitating the development of widely accessible stewardship methods.
- The misuse and overuse of antibacterials adversely affect patients admitted to intensive care units (ICUs).
- Further research is urgently required to determine the most effective ways to implement ASPs in LMICs.

**What this study adds:**

- By establishing a quality improvement collaborative (QIC), we showcased an improvement in antibacterial utilization within ICUs in a low- to middle-income country.
- Additionally, a reduction in healthcare-associated infections is evident.
- Moreover, the QIC effectively strengthened the capabilities of infection control and prevention in participating ICUs.

**How this study might affect research, practice, or policy:**

- This study is among the initial endeavors in a middle-income country to evaluate the efficacy and essential strategies for establishing antimicrobial stewardship programs.
- This study could serve as a foundational reference for upcoming teams aiming to introduce similar programs in the region.

## Background

Health care-associated infections (HAI), a global health care problem, are more prevalent in low- and middle-income countries (LMICs)[1]. The frequency of patient intensive care unit-acquired infections is nearly three times higher than that in high-income countries (42.7 episodes per 1,000 days), and antimicrobial resistance (AMR) is also higher in LMICs[1]. AMR is a rapidly worsening global problem. The overuse and misuse of antibacterials, poor sanitation, low vaccination rates, and poor infection prevention and control practices all contribute to the high rate of drug-resistant infections in LMICs[2].

In comparison with infections caused by susceptible bacteria, those caused by multidrug-resistant organisms (MDRO) are associated with a higher incidence of mortality and prolonged hospital stay[3]. Unexposed patients can be adversely affected by the spread of MDRO and *C. difficile*[4]. Serious adverse events occur in approximately 20 percent of hospitalized patients receiving antibacterials[5].

Antimicrobial Stewardship Programs (ASP) have been developed to optimize the treatment of infections, reduce infection-related morbidity and mortality, limit the appearance of MDRO, and reduce unnecessary antimicrobial use. This practice ensures optimal selection, dose and duration of antimicrobials, and leads to the best clinical outcome for the treatment or prevention of infection[6]. Hospital ASP can increase infection cure rates while reducing treatment failures, *C. difficile* infections, adverse effects, antibacterial resistance, hospital costs, and length of stay[7–9].

Patients admitted to ICUs are commonly treated with one or more antimicrobial agents during their stay, and different international and national registries of antimicrobial use in ICU patients have been developed in order to optimize the antimicrobial use and consequently reduce MDRO [10]. During the first part of the COVID-19 pandemic an increase in the consumption of antibacterials was observed[11], and nearly three-quarters of patients received antibacterials therapy[12]. Bacterial co-infection rates for SARS-CoV-2 have been estimated between 6.1% and 8.0%. Antibacterials prescription was significantly higher than the prevalence of bacterial co-infection, suggesting that many antimicrobial prescriptions were unnecessary, increasing the risk of preventable harm, including adverse events, *C. difficile* infections, and AMR. However, developing countries may go through periods of deficient healthcare conditions that affect the availability of antimicrobial agents. Under these conditions, a reduction in antimicrobial consumption may not represent a favorable outcome[13].

Given that the Centers for Disease Control and Prevention suggest that nearly 50% of antimicrobial use is unnecessary or inappropriate[14], widely available methods of stewardship need to be developed. The challenges in antimicrobial stewardship in both high-income countries and LMICs are significant. These include prescribing broader-spectrum antibiotics due to fear of not covering a specific pathogen with narrow-spectrum drugs, and unnecessarily escalating treatment in response to new clinical events such as fever or hypotension within a few hours of treatment initiation[15]. To date, there is little consolidated evidence regarding the effectiveness of ASPs in LMICs[16,17]. Introducing ASPs in LMICs poses challenges owing to factors such as limited availability and access to antimicrobials, lack of diagnostics technologies, and poor adherence to treatment[16]. Further research is urgently required to determine the most effective ways to implement ASPs in LMICs, without compromising the quality of care provided to patients[18].

To address gaps in performance, quality improvement collaboratives (QIC) have been used to improve health care for several decades disseminating evidence and learnings from implementation science[19–21]. The QIC proved to be useful and effective in producing rapid changes at the scale.

Our aim was to support ICUs by implementing a multi-faceted intervention, composed of a range of antimicrobial stewardship strategies intended to enhance the quality of antibacterials prescription by reducing its overuse and increasing the use of narrow-spectrum agents (de-escalation). Our specific objectives were directed to ICU patients and were to reduce antibacterial days of therapy, the defined daily dose of antibacterials, HAIs associated with multidrug resistant bacteria, and HAIs associated with C. difficile.

## Methods

### Study design

We developed a QIC preceded by a formative phase, enrolling ten ICUs over an eleven-month period under the design of an uncontrolled interrupted time series with a baseline period (BP) of 16 weeks and an implementation period (IP) of 32 weeks. This study was conceived by applying the Breakthrough Innovative Series for QIC from the Institute for Healthcare Improvement. It involves the use of healthcare teams from different sites to improve performance on a specific topic by collecting data and testing ideas with plan-do-study-act (PDSA) cycles supported by coaching and learning sessions[22,23]. The QIC was supported by the concept that networks of facilities can be harnessed into learning systems that accelerate improvement in healthcare performance with the potential to achieve results on a large scale.

### Settings

Based on the eligibility criteria, ten ICUs in the public sector of the national health system were recruited. Inclusion criteria for ICUs encompassed belonging to the public sector, having an implemented infection control program, having at least prevalence measurements of DOT or DDD, unit size above eight beds in regular situations (not expanded by the pandemic) and having signed a letter of commitment from hospital and ICU leaders to assure intervention deployment and data collection throughout the project. The exclusion criterion was HAI outbreak.

### Participants

Healthcare workers (HCW) at participating ICUs were the targets of the intervention and research subjects of this study. Primary and secondary outcomes were measured in patients admitted to the participating ICUs. The inclusion criteria for HCW were physicians, pharmacists, and nurses at participant ICUs. The inclusion criterion for patients was having received antibacterials during ICU admission in the participating ICUs. Patients were excluded if they had received antibacterials for prophylaxis, abdominal infections, tuberculosis, VIH, or febrile neutropenia owing to cancer.

### Formative research

We conducted a qualitative study involving 19 remote individual semi-structured interviews with healthcare professionals working in the ICU of the participating hospitals in the collaborative study. The interviews took place between June and July 2022, and were conducted via phone or remote means and audio recording. Data analysis was conducted using a theory-guided approach (Normalization Process Theory - NPT)[24]. The qualitative data management software Atlas.ti v8.4 was utilized to facilitate the coding process of the data.

### Intervention

We used a comprehensive implementation science-based package. The implementation of the intervention can be found in the Template for Intervention Description and Replication (Table 1), and the theory of change is outlined in the driver diagram (Figure 1)

### Measures

We designed a study with two phases: an initial phase involving 8 biweekly measurements followed by an intervention, and a subsequent phase with 16 biweekly measurements. The primary outcome measures were the number of antibacterial days of therapy (DOT) per 1000 in-patient days and the defined daily dose (DDD) of antibacterial per 1000 in-patient days. DDD measures drugs administered as multiples of the assumed average maintenance dose per day for a specific patient[25]. DOT is the number of days of antibacterial therapy administered to a patient, regardless of the number of doses administered or dosage strength[25]. Secondary outcome measures included: 1) de-escalation, which is the proportion of empirical therapy changed to pathogen-directed therapy as soon as culture results became available; 2) rates of healthcare-associated infections (HAIs) associated with multidrug-resistant bacteria, including ventilator-associated pneumonia (VAP), central line-associated bloodstream infections (CLABSI), and catheter-associated urinary tract infection (CAUTI); 3) rates of HAIs associated with *C. difficile*, not present before admission; 4) length of stay in days (truncated at day 28 of ICU stay); and 6) ICU mortality or mortality before day 28 of ICU stay.

**Figure 1.**
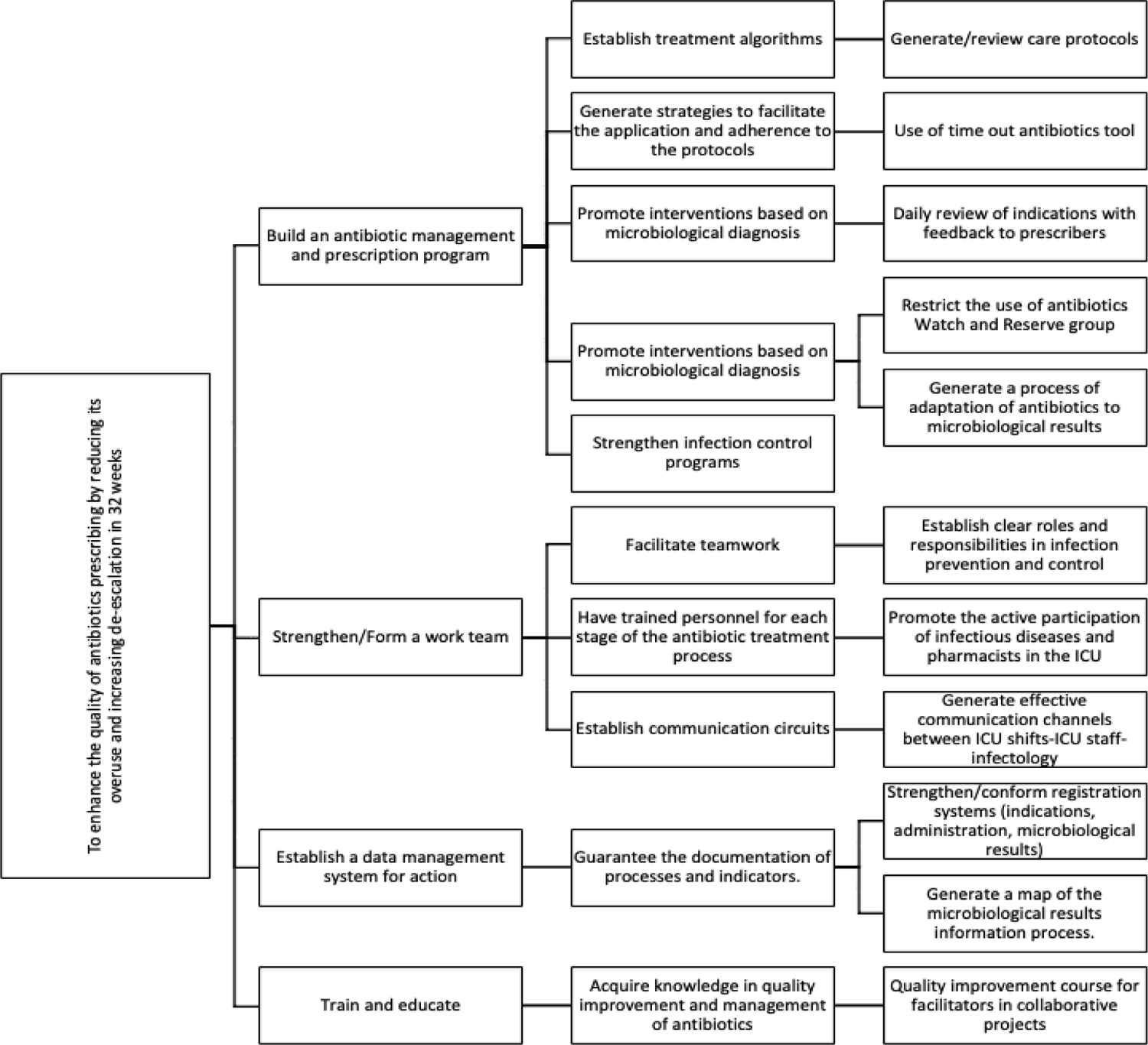
Theory of change driver diagram

**Table 1.**
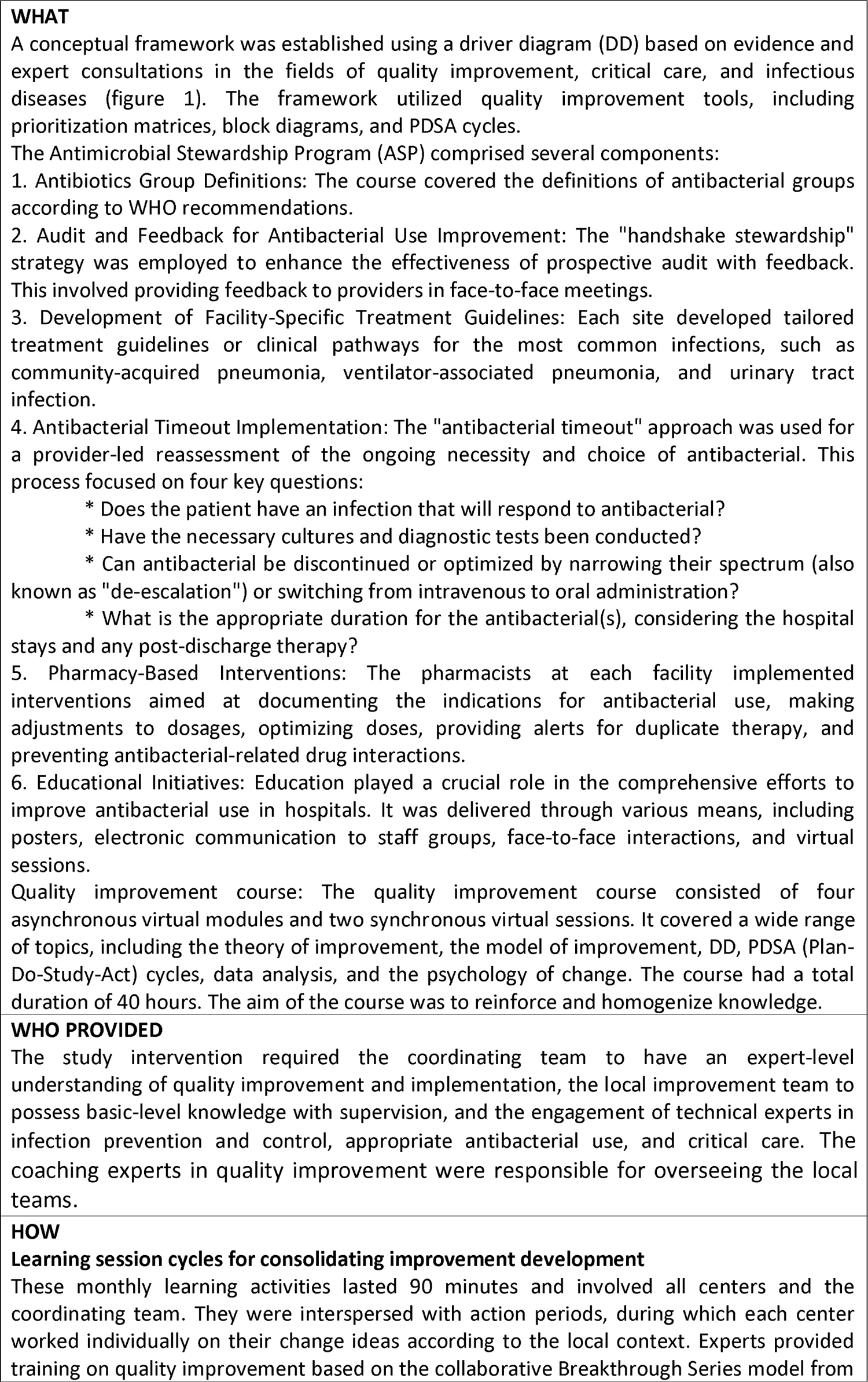
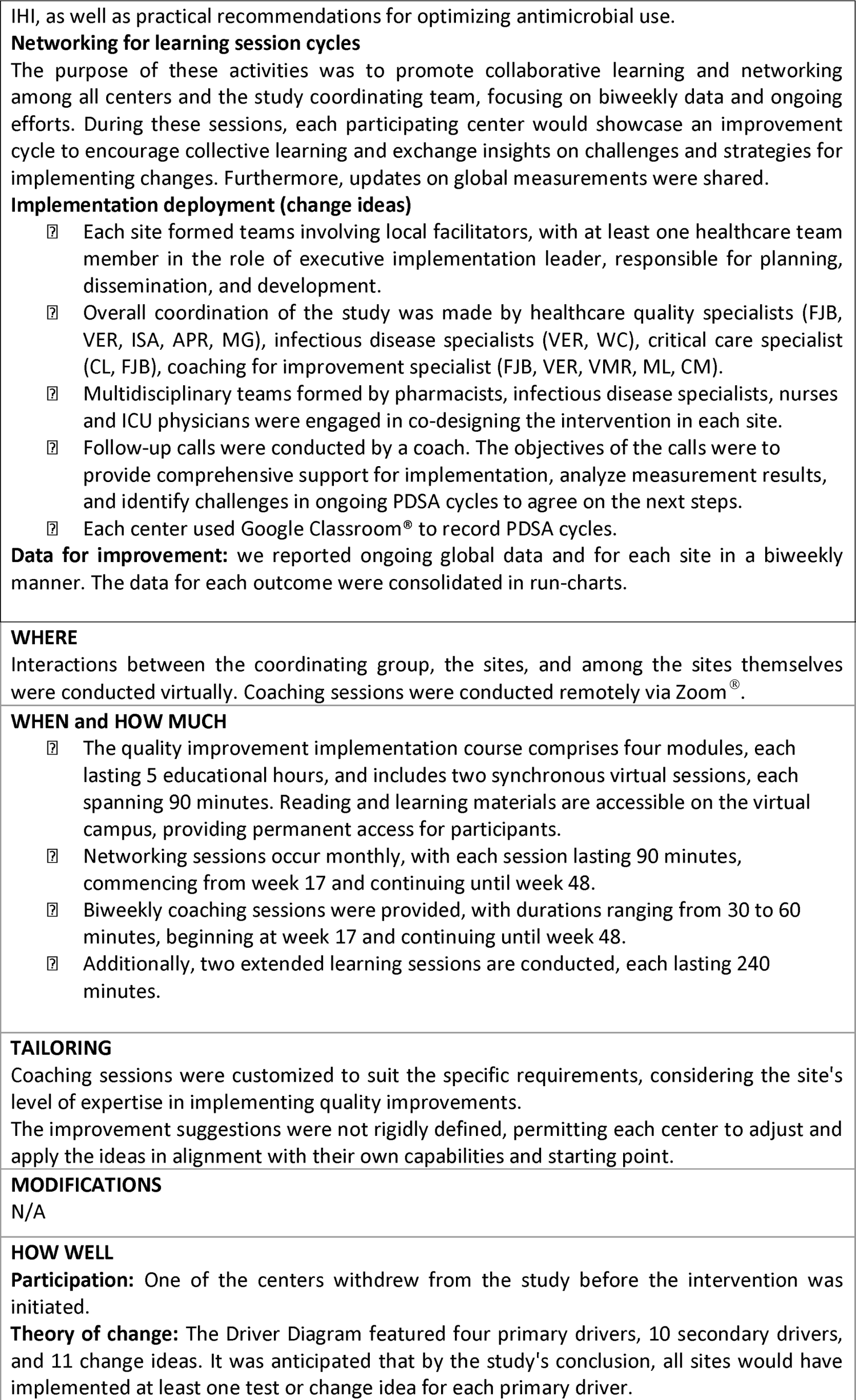
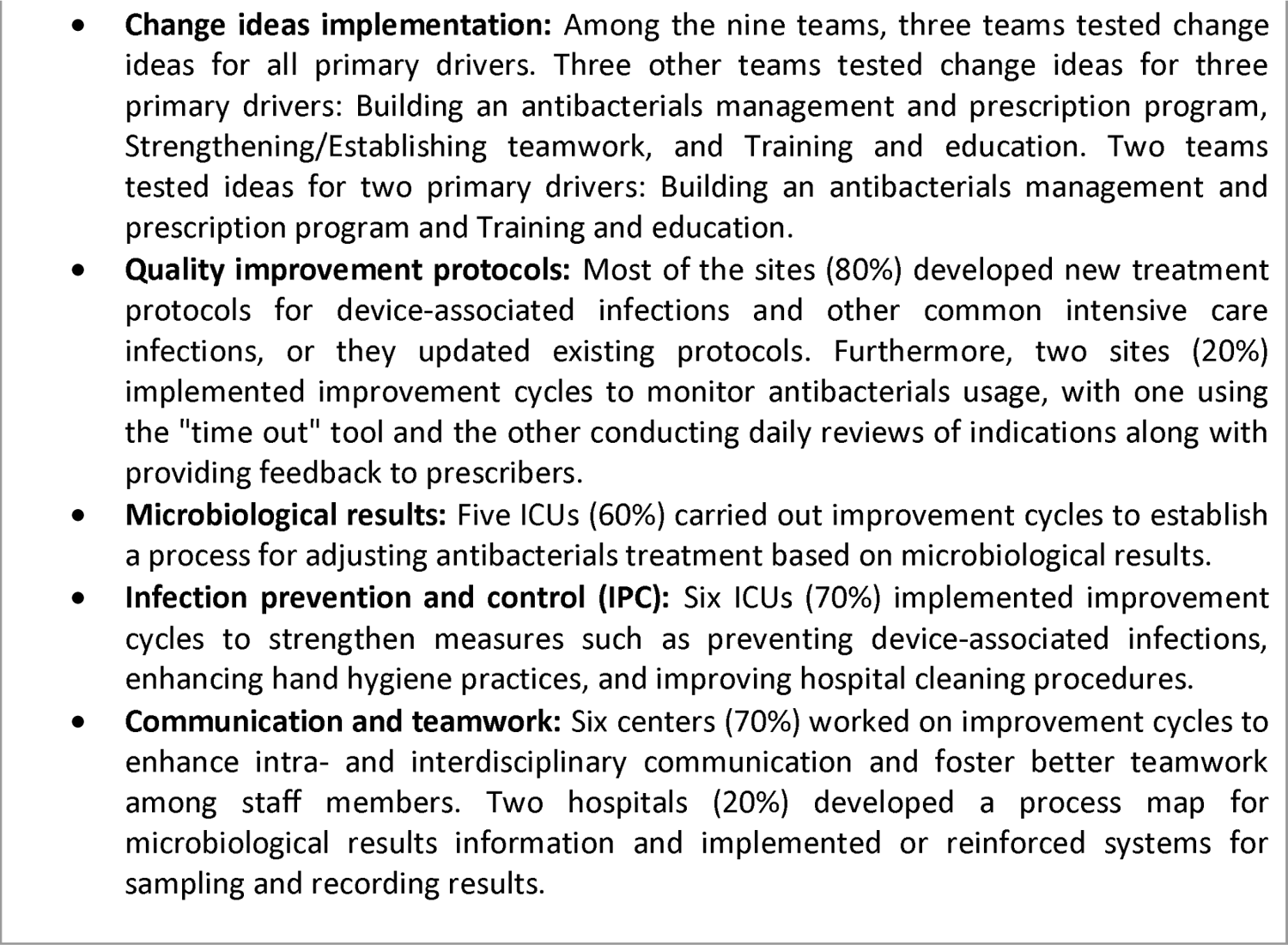
The Template for Intervention Description and Replication.

We measured other variables, such as the Acute Physiology and Chronic Health Disease Classification System II (APACHE II) score, Sequential Organ Failure Assessment (SOFA) score, age, sex, and invasive procedures (mechanical ventilation, central venous catheter, urethral catheter, extrarenal depuration techniques, ventricular drainage catheter, and total parenteral nutrition).

Additionally, we assessed the Infection Prevention and Control (IPC) Assessment Framework at both the beginning and end of the study[26]. This framework was designed to support the implementation of ASP in healthcare facilities. Its objective is to evaluate current IPC activities and resources within facilities and identify strengths and areas for improvement. The assessment serves as a guide for future planning, helping organizations optimize their IPC practices and allocate resources effectively.

### Data collection

Data were extracted from health records by unit data collectors and were gathered in a Redcap® form. In addition, each team reported on the development of improvement opportunities using a standardized report developed for this purpose. Virtual follow-up visits (audits) were conducted in all participating units, and continuous communication was maintained between site coordinators and data collectors via telephone, email, and WhatsApp®.

The IECS data unit supervised database monitoring, consolidation, and analysis as necessary, utilizing Redcap® and adhering to Good Clinical Research Practices. Regular assessments of compliance with the project protocols were conducted, including unit screening, data collection, adherence to the intervention protocol, and data quality.

### Statistical Analysis

The participating ICUs and their patients were characterized for the baseline and implementation periods. Absolute and relative frequencies were reported for categorical variables, while the median and interquartile range (IQR, quartiles 1 - 3) were presented for continuous variables because of their non-normal distribution. For comparison, the chi-square test was used for categorical variables, and the Wilcoxon test was used for continuous variables.

The DOT and DDD outcomes were presented as rates per 1,000 hospitalization days and calculated for both periods. The effect of the intervention was measured using the ratio of rates: Rate IP/Rate BP. To estimate this ratio, a generalized linear model, assuming a negative binomial distribution, was employed. To estimate the DOT, we used a model in which the outcome was the number of antibacterial days of therapy, the offset variable was the number of intensive care unit days of stay. In the case of the DDD, the outcome was the daily dose of antibacterial and the same offset variable. In the models, the variable of interest in the models was a variable that represented the period. Both the crude estimated ratio of the rates and the adjusted ratio of the rates by SOFA score, septic shock, and kidney failure were reported. Additionally, the impact of the intervention was analyzed across subgroups by considering the following variables: SOFA score (high vs. low), APACHE II score (high vs. low), renal failure (yes vs. no) and septic shock (yes vs. no).

The adequacy of treatment and distribution of antibacterial, including median doses, were reported for each period. Furthermore, potential differences in the characteristics of microbiological rescues between the two periods were evaluated using the chi-squared test. Microorganism type, time from sample collection to microbiological rescue, antibacterial resistance, type of resistance, need for antibacterial adjustment, and the time from microbiological rescue to antibacterial dispensation were documented for both periods. Other outcomes were compared using the same method as that used for the primary outcomes. Data were analyzed using R version 4.0.2 (The R Foundation).

### Ethics

Institutional Review Board approval was obtained for all ICU participants. Informed consent was obtained from all the participants.

## Results

### Formative research

Work overload was identified as a significant barrier to any activity. Participants emphasized that the intervention activities had to be simple, brief, and asynchronous to facilitate adherence. All participants highlighted the importance of incorporating specialists in infectious diseases, pharmacy, and infection control into intensive care teams. The importance of constant follow-up and education, as well as the elimination of the administrative burden, were underlined for the success of the intervention. It was recommended that nursing teams be supported and monitored as they were identified as crucial for the intervention, although with different types of flaws.

Ensuring the monitoring of data load updates was crucial, as some sites faced a shortage of human resources or lacked computerized medical records. It was essential to offer specialized support to teams that did not have their own infectious disease specialists. Teams that reported financial barriers to accessing specific medications or experienced significant delays in receiving laboratory test results were expected to encounter specific problems.

Participants explained that the study introduction module would be beneficial for all participants. They suggested providing concise information with access to comprehensive resources for the medical teams on duty. Additionally, they recommended tailoring training workshops to ensure the facilitators’ availability. Suggestions were made to record workshops to facilitate attendance, while emphasizing the importance of including synchronous or in-person sessions for more personalized training and follow-up.

### Outcomes results

We recruited 912 patients during the study period: 357 during BP (between 3/28/2022 and 7/17/2022) and 555 during IP (between 7/18/2022 and 2/26/2023). The patients in the IP group had more severe disease, with higher APACHE II score, higher SOFA score, higher prevalence of kidney failure, and higher mortality rate in the ICU. However, the standardized mortality ratio (SMR) by the APACHE II score between study groups was similar (BP group 0.99 (0.81-1.21) vs. IP group 1.17 (1.01- 1.35), ratio of the SMR 1.18 (0.92;1.51); p=0.185). The complete characteristics of the patients and type of discharge in both periods are shown in Table 2.

**Table 2.** Patients’ characteristics and type of discharge between study phases.

| Characteristic | Baseline<br>N= 357<br>n/N (%) | Implementation<br>N= 555<br>n/N (%) | p-value** |
| --- | --- | --- | --- |
| Days of hospitalization in ICU* | 10 (5, 20) | 9 (4, 19) | 0.679 |
| Sex |  |  | 0.612 |
| Male | 214/357 (59.9%) | 342/555 (61.6%) |  |
| Female | 143/357 (40.1%) | 213/555 (38.4%) |  |
| Age* | 51 (34, 64) | 50 (36, 65) | 0.439 |
| Weight* | 75 (65, 90) | 75 (70, 85) | 0.871 |
| APACHE II Score* | 15 (11, 20) | 17 (12, 21) | 0.036 |
| High Score APACHE II<sup>α</sup> | 152/357 (42.6%) | 284/555 (51.2%) | 0.011 |
| SOFA<sub>24</sub> Score* | 5 (3, 8) | 6 (4, 9) | 0.006 |
| High Score SOFA<sup>β</sup> | 170/357 (47.6%) | 318/555 (57.3%) | 0.004 |
| Kidney failure** | 118/357 (33.1%) | 231/555 (41.6%) | 0.009 |
| Use of mechanical ventilation | 270/357 (75.6%) | 420/555 (75.7%) | 0.988 |
| Days of Mechanical Ventilation* | 10 (4, 18) | 9 (4, 18) | 0.974 |
| Days free of Mechanical Ventilation*<sup>Ω</sup> | 5 (0, 20) | 0 (0, 19) | 0.018 |
| Use of central venous catheter | 311/357 (87.1%) | 487/555 (87.7%) | 0.778 |
| Days of use of Central Venous Catheter* | 10 (5, 17) | 9 (5, 16) | 0.886 |
| Use of vesical catheter | 327/357 (91.6%) | 525/555 (94.6%) | 0.075 |
| Days of use of vesical catheter* | 10 (4, 20) | 9 (5, 18) | 0.787 |
| Use of extra-renal clearance techniques | 39/357 (10.9%) | 74/555 (13.3%) | 0.281 |
| Days of use of extra-renal clearance techniques* | 5 (3, 16) | 6 (3, 10) | 0.796 |
| Use of ventricular drainage catheter | 13/357 (3.6%) | 29/555 (5.2%) | 0.265 |
| Days of use of ventricular drainage catheter* | 8 (4, 12) | 3 (3, 9) | 0.144 |
| Use of parenteral nutrition | 14/357 (3.9%) | 11/554 (2.0%) | 0.081 |
| Days of use of parenteral nutrition* | 9 (7, 14) | 12 (8, 14) | 0.621 |
| Type of discharge |  |  | 0.018 |
| Hospitalization room | 200/357 (56.0%) | 282/555 (50.8%) |  |
| Death | 94/357 (26.3%) | 189/555 (34.1%) |  |
| Other institution/third level | 26/357 (7.3%) | 23/555 (4.1%) |  |
| Home Hospitalization | 4/357 (1.1%) | 2/555 (0.4%) |  |
| Remains in ICU on day 28 | 33/357 (9.2%) | 59/555 (10.6%) |  |

\*Median (IQR) reported
\*\* P-value obtained from Wilcoxon test was used or Chi- square test was used.
<sup>α</sup> High Score APACHE II corresponds to values greater than 16.
<sup>β</sup> High Score SOFA<sub>24</sub> corresponds to values greater than 5.
<sup>Ω</sup> Calculated as 28 days - days of mechanical ventilation.

The distribution of illness severity differed between periods. The occurrence of sepsis (31.6% in the BP group and 36.1% in the IP group and septic shock (33.8% in the BP group and 40.0% in the IP group) was higher in the IP group (p<0.001). The site of infection was similar between the phases of the study, with nearly 40% of respiratory infections and 20% of VAP (see eTable 1, supplement).

We did not detect any changes in DOT between the phases (BP 1,133 and IP 1,112, RR 0.98 (0.95-1.02); p=0.297). Even after adjusting for illness severity factors (APACHE II score >16, SOFA score >5, septic shock, and renal failure), no significant differences were found (RR 0.97 (0.90; 1.04), p=0.355). However, we observed a decrease in antibacterial DDD during the implementation phase (BP: 1,301; IP: 1,193, RR 0.92 (0.89, 0.95); p<0.001). This difference in DDD between the periods remained evident after adjusting for illness severity (RR 0.87 (0.80; 0.96), p=0.004). The DOT and DDD stratified analysis for illness severity is shown elsewhere (see eTables 2 and 3, supplement). The distribution of antibacterial, including median doses, were reported for each period (see eTable 4).

Approximately half of the treatments necessitated antibacterial adjustments following microbiological results, comprising 55.5% of the BP group and 49.7% of the IP group, (p=0.069). De-escalation, defined as modifying the antibacterial within the initial 24 hours after microbiological intervention, occurred more frequently in the IP, with 181 out of 292 cases (62.0%) than in 110 out of 243 cases (45.3%) in the BP, indicating a significant disparity (p<0.001). The different ranges from microbiological rescue to antibiotic prescription are shown in eTable 5, supplement.

No notable disparities in antibacterial resistance were observed between the two periods (BP 158/438 (36.1%) and IP 210/587 (35.8%); p=0.999). However, methicillin resistance was more prevalent in the IP group. Moreover, there were no discernible differences in the resistance patterns regarding the type of carbapenem resistance (see eTable 6, supplement). Despite the low percentage of antibacterial that switched to oral administration, a higher frequency was observed during IP (1.6% in the IP group compared to 0.5% in the BP group). Throughout the study, we monitored the HAIs caused by MDROs. We noticed a reduced rate of VAP and CAUTI in the IP group, while observing no alterations in CLABSI and HAIs linked to *C. difficile* (see Table 3).

**Table 3.**
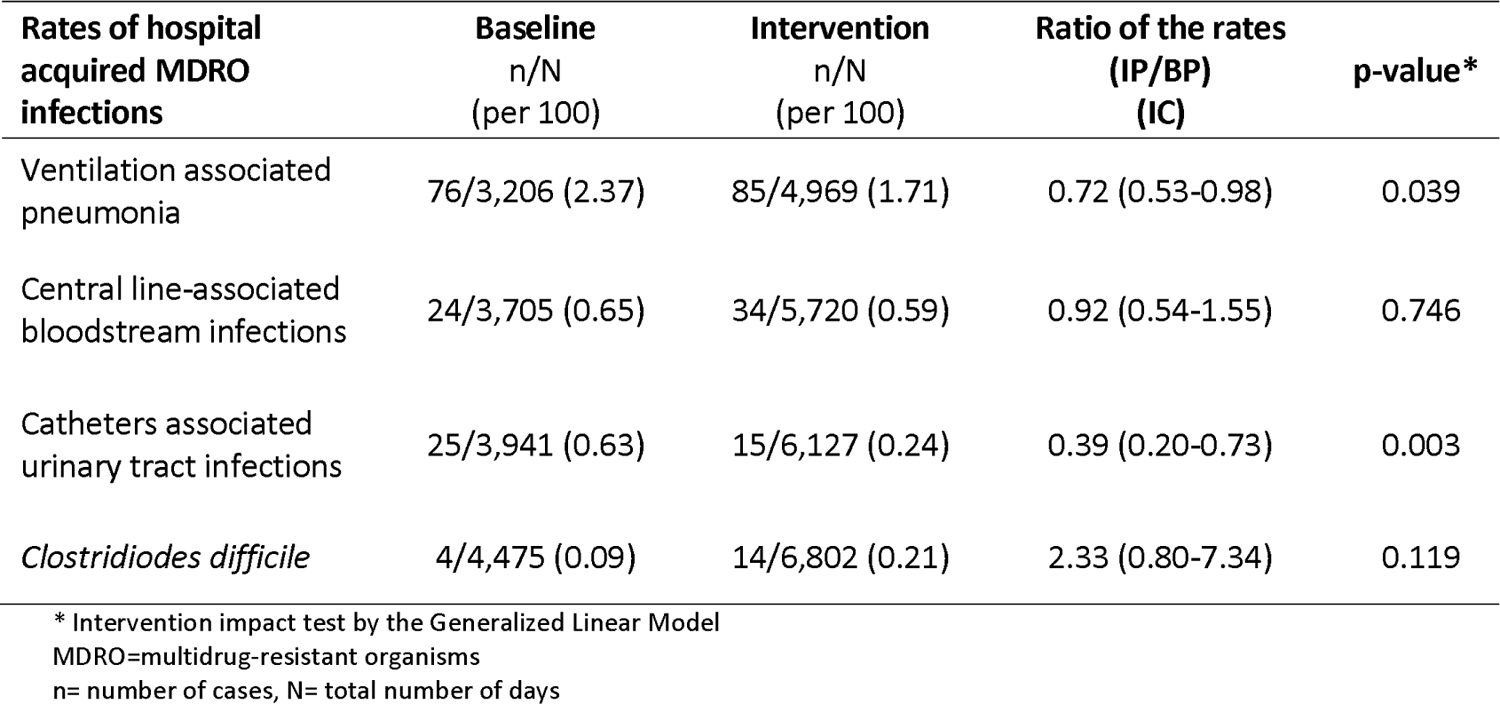
Muti-resistant microorganism healthcare associated infections rate between study periods.

| Rates of hospital acquired MDRO infections | Baseline<br>n/N<br>(per 100) | Intervention<br>n/N<br>(per 100) | Ratio of the rates<br>(IP/BP)<br>(IC) | p-value* |
| --- | --- | --- | --- | --- |
| Ventilation associated pneumonia | 76/3,206 (2.37) | 85/4,969 (1.71) | 0.72 (0.53-0.98) | 0.039 |
| Central line-associated bloodstream infections | 24/3,705 (0.65) | 34/5,720 (0.59) | 0.92 (0.54-1.55) | 0.746 |
| Catheters associated urinary tract infections | 25/3,941 (0.63) | 15/6,127 (0.24) | 0.39 (0.20-0.73) | 0.003 |
| *Clostridioides difficile* | 4/4,475 (0.09) | 14/6,802 (0.21) | 2.33 (0.80-7.34) | 0.119 |

\* Intervention impact test by the Generalized Linear Model MDRO=multidrug-resistant organisms n= number of cases, N= total number of days

There was a marginal and non-significant shift in the percentage of readmissions within 48 hours observed between the study periods. In the BP group, 3 out of 357 cases (0.8%) resulted in readmission, whereas in the IP group, 2 out of 555 cases (0.4%) led to readmission (p=0.400).

We noted an enhancement in The Infection Prevention and Control Assessment Framework in comparison to the initial assessment[26]. The baseline score was 417 points, while post-implementation, the score increased to 480 points. Both periods showed an intermediate IPC level. Remarkably, six out of the eight dimensions displayed improvement post-intervention, signifying positive advancement in these areas (see figure 2).

**Figure 2.**
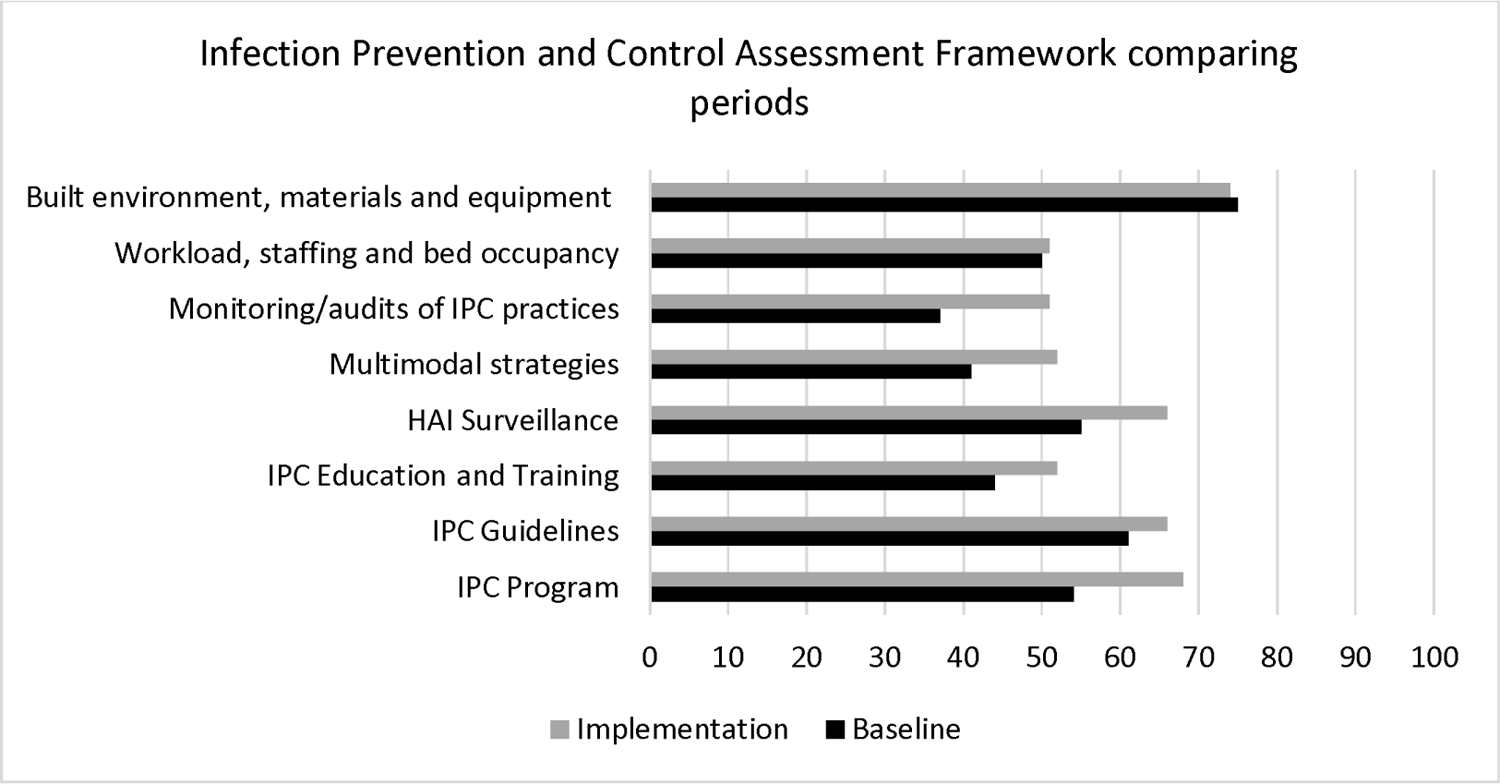
Infection Prevention and Control (IPC) Assessment Framework score comparing periods. Higher scores show a better behavior in the different dimensions of IPC.

## Discussion

Our key finding underscores the success of a multifaceted intervention centered on optimizing antimicrobial usage. This initiative resulted in a decrease in DDD of antibacterial agents, an improvement in the de-escalation process towards narrow-spectrum drugs, and a reduction in HAIs linked to MDROs, despite having limited resources. Moreover, we successfully improved the IPC score compared to the initial baseline measurements. Nevertheless, we were unable to decrease the DOT during the IP.

In addition to these findings, crucial data regarding the profile of MDROs was reported by measuring their incidence rate ratio over a period of 10 months. This data is not widely available in Latin American countries. In a recent study covering 333 ICUs across 52 European countries with 2600 patients analyzed, known as the EUROBACT-2 study, a comparable profile with our study of Difficult-to-Treat Resistance, primary sources of HAIs, and the categorization of microorganisms was identified[27]. This study included mainly ICUs within the public sector and standardized patients based on the severity of illness, like our approach. The observed mortality rate was high, like ours, possibly linked to Difficult-to-Treat Resistance[28]. Adequate antimicrobial therapy was administered to 52% of patients within 24 hours of blood culture sampling[27]. In our study, we initially observed a similar rate of adequate antimicrobial therapy during the baseline phase. However, following the IP, we witnessed a significant improvement in the adequacy of treatment.

The primary objective of this study was to optimize antimicrobial usage, focused on antibacterials, by employing a method that ensured an appropriate dose and treatment duration, while also avoiding unnecessary prescriptions and discontinuing treatment when deemed adequate. In addition, infection prevention strategies such as proper hospital and hand hygiene, as well as the appropriate use and management of medical devices (e.g., probes, catheters, and respirators), were considered. The application of shorter antimicrobial courses should not be approached uniformly, but rather on an individualized basis[29]. Guidelines for antimicrobial treatment offer valuable resources for determining when to opt for a shorter treatment duration, considering factors such as source control, biomarkers (when available), antimicrobial susceptibility, and the severity of the infection. Additionally, further research on shortening treatment duration, including both observational and clinical trials, would be beneficial in developing a more objective and tailored approach to this issue[15].

The results of this study suggest that the resistance to antibacterial was not diminished, and there was a change to higher methicillin resistance and a lower but not significant vancomycin resistance. It is important to note that most stewardship programs and research studies on this topic have shown an association with 30% in antibacterial consumption rates and a 10% reduction in antibacterial prescriptions[18]. Most of the studies have not measured the impact on microbiology[30,31]. The implications of implementing ASP in resource-constrained environments continue to be a subject of uncertainty, with the potential outcomes remaining indeterminate[18].

Our study notably enhanced IPC capacity, particularly in dimensions unrelated to infrastructure, workload, staffing, and bed occupancy. The intervention did not aim to augment equipment and material acquisition, nor was it intended to alter workload due to a low budgeted intervention. Significantly, the dimensions showing the most improvement were the development of IPC programs and monitoring/auditing of IPC practices. This aligns with our theory of change (Figure 1), which includes secondary drivers aimed at reinforcing IPC programs. Approximately 70% of the ICUs employed the secondary driver by implementing PDSA cycles to improve hand hygiene and prevent VAP, CAUTI, and CLABSI.

The intervention was modified in response to the findings of our formative research, integrating experts in infectious diseases, pharmacy, and infection control into the intensive care teams. Additionally, we created an educational module aimed at enhancing quality improvement capabilities, which was disseminated via virtual synchronous sessions and a virtual campus. However, we were unable to address the issue of staff workload and material resources, as these factors were beyond the scope and budget of our intervention.

### Strengths and limitations

The intervention, as depicted in the project’s driver diagram, incorporated multidisciplinary activities, and considered human factors such as team building, role definition, effective communication, and continuous improvement training. The implementation of multimodal initiatives, which were co-designed by operative professionals with a human-centered approach, proved effective in managing and sustaining change. This framework aligns with the principles of the Improvement Breakthrough Series of the Institute of Healthcare Improvement[22]. Analysis of the intervention and implementation revealed the need for adaptation to each center’s resources and structure.

Despite the limited resources, the data collection was of exceptional quality and provided highly valuable information pertaining to the type of antibacterial drug utilized, the respective dosages and duration of usage, the culture type, and the time to rescue, organism resistance patterns, as well as de-escalation practices.

Certain elements of the intervention necessitated deep behavioral modifications among staff members, and in some cases, the institutional culture was not mature enough to facilitate these changes. Although the teams highly valued the implemented changes. Additionally, forming multidisciplinary teams posed challenges in some centers, resulting in delays in the participation of the designated “change agent” responsible for supporting and guiding the implementation. With a longer intervention period, the gradual changes that began to occur would have been more noticeable in the results. Moreover, the timing of the intervention, which coincided with the end of the year and the vacations of a significant number of the personnel, could had been a factor that weakened the implementation at a crucial stage when changes tended to solidify, thus limiting its effect. The participating sites varied in context and quality improvement, requiring continual support, and coaching for most sites. This highlights the need for an extended exposure period, particularly in low- or middle-income regions, where healthcare quality lags for high-income countries.

Regarding the improvement cycles conducted in the centers, developing treatment guidelines requires substantial effort and time. This involved conducting a literature review, assessing the frequently isolated pathogens in the unit, and preparing and disseminating new guidelines. These activities sometimes resulted delayed subsequent improvement cycles. Similarly, after the guidelines were created, it took time for the team to incorporate the recommendations into daily practice, potentially delaying their impact on results. Staff members, particularly in critical patient cases, often question the prescription of antimicrobials with a lower spectrum or dose, despite supporting evidence. We encountered challenges in assessing adherence to guidelines, as the ICUs in the BP lacked guidelines or utilized outdated ones.

Furthermore, coordination and monitoring of the entire project was conducted entirely through virtual means by the project coordination team. Poor connectivity and the lack of on-site coaching and monitoring for sites with less experienced professionals in quality improvement, antibacterial stewardship, and IPC have made it challenging to build strong and consistent teams. These sites require stronger support to sustain the improvement cycles.

To our knowledge, this is one of the first studies to assess the effectiveness and necessary strategies for implementing antimicrobial stewardship programs in low-income and middle-income countries. This could serve as a starting point for future teams seeking to implement similar programs in the region. Moreover, the study enhanced the capacity for infection control and prevention in participating intensive care units.

## Conclusions

By executing an ASP through a QIC, we effectively enhanced antibacterial use in ICUs across Argentina. Furthermore, we noticed improvements in IPC metrics and overall capabilities for quality enhancement. Nevertheless, it is crucial to recognize that achieving a notable reduction in DOT might necessitate further time and effort for ASP implementation. Improving the utilization of antibacterial treatments may help to minimize infections caused by MDRO. The brief duration of the intervention in this study may have impeded the ability to detect substantial reductions in antimicrobial resistance and the desired enhancements in antimicrobial utilization. Based on the findings, it is speculated that a more extended intervention period and a stronger integration of antibacterial stewardship, device management, and infection prevention strategies are essential to achieve a substantial impact on microbiology.

Supplementary matherial

## Data Availability

https://osf.io/5v7xa/?view_only=111e421428c5463385190685e6fa1cca

## Notes

### Competing Interest Statement

The authors have declared no competing interest.

### Funding Statement

PFIZER COMPETITIVE GRANT PROGRAM ID: 68339261

### Author Declarations

Approvals and dates from Institutional Review Board of Ministry of Health of Neuquen - Hospital Castro Rendon (05/07/2022), Institutional Review Board of Hospital de infecciosas F. J. Muniz (11/15/2021), Institutional Review Board of province of Jujuy (12/22/2021), Institutional Review Board of the Hospital Ignacio Pirovano (11/10/2021), Institutional Review Board of the Hospital General de Agudos Bernardino Rivadavia (01/10/2022), Institutional Review Board of HOSPITAL SAN MARTIN DE LA PLATA (11/25/2021), Institutional Review Board of Hospital Francisco Lopez Lima (11/20/2021), Institutional Review Board of Hospital Simplemente Evita (04/07/2022), Institutional Review Board of Hospital Evita de Lanus (01/20/2022)

