## Supplementary matherial for "Addressing the aftermath of the COVID-19 pandemic: A quality improvement collaborative to optimize the use of antibacterials in Argentine Intensive Care Units"

### Supplementary appendix

*eTable 1. Severity of clinical presentation and site of infection origin between study phases.*

| Characteristic | Baseline (N = 547) N (%) | Implementation (N = 812) N (%) | P-value* |
|---|---|---|---|
| **Severity of infection** | | | <0.001 |
| Sepsis | 173 (31.6%) | 293 (36.1%) | |
| Septic shock | 185 (33.8%) | 325 (40.0%) | |
| Infection without sepsis | 189 (34.6%) | 194 (23.9%) | |
| **Site of infection** | | | 0.083 |
| Respiratory infection | 229 (41.9%) | 321 (39.5%) | |
| Urinary tract infection | 37 (6.8%) | 49 (6.0%) | |
| CNS infection | 18 (3.3%) | 46 (5.7%) | |
| Skin and soft tissue infection | 45 (8.2%) | 74 (9.1%) | |
| Ventilator-associated pneumonia | 114 (20.8%) | 149 (18.3%) | |
| Central venous catheter associated infection | 40 (7.3%) | 55 (6.8%) | |
| Urinary catheter-associated infection | 22 (4.0%) | 21 (2.6%) | |
| C. difficile colitis | 4 (0.7%) | 15 (1.8%) | |
| Primary bacteremia | 28 (5.1%) | 55 (6.8%) | |
| Other | 10 (1.8%) | 27 (3.3%) | |

*Chi-squared test

*eTable 2. Illness severity stratified analysis for days of antibacterial treatment.*

| Strata | Baseline period | Implementation period | Ratio of the rates |
|---|---|---|---|
| | n/N (per 1,000) | n/N (per 1,000) | (Post/Pre) (IC 95%)Ω |
| **Sofa score**[1] | | | |
| High (n = 488) | 2754 / 2382 (1156.2) | 4478 / 4114 (1088.5) | 0.97 (0.88;1.08) |
| Low (n = 424) | 2318 / 2093 (1107.5) | 3087 / 2688 (1148.4) | 1.00 (0.89;1.12) |
| **Apache score**[2] | | | |
| High (n = 436) | 2513 / 2177 (1154.3) | 4123 / 3633 (1134.9) | 0.94 (0.85;1.04) |
| Low (n = 476) | 2559 / 2298 (1113.6) | 3442 / 3169 (1086.2) | 0.98 (0.93;1.03) |
| **Kidney failure** | | | |
| Yes (n = 349) | 2130 / 1637 (1301.2) | 3380 / 2717 (1244.0) | 0.99 (0.88;1.11) |
| No (n = 563) | 2942 / 2838 (1036.7) | 4185 / 4085 (1024.5) | 0.95 (0.86;1.06) |
| **Septic shock** | | | |
| Yes (n = 389) | 2484 / 1895 (1310.8) | 4157 / 3350 (1240.9) | 0.98 (0.88;1.09) |
| No (n = 523) | 2588 / 2580 (1003.1) | 3408 / 3452 (987.3) | 0.95 (0.86;1.06) |

Ω Intervention impact tested by the Generalized Linear Model
[1] High SOFA score: scores > 5, Low SOFA score: scores <=5
[2] High APACHE score: scores >16, Low APACHE score: scores <=16
n: number of days of antibacterial treatment, N: number of intensive care unit days of stay

*eTable 3. Illness severity stratified analysis for antibacterial defined daily doses.*

| Strata | Baseline period n/N (per 1,000) | Implementation period n/N (per 1,000) | Ratio of the rates (Post/Pre) (IC 95%) Ω |
|---|---|---|---|
| **$SOFA_{24}$ score**[1] | | | |
| High (n = 488) | 3264 / 2382 (1370.4) | 4776 / 4114 (1161.0) | 0.86 (0.76; 0.97) |
| Low (n = 424) | 2558 / 2093 (1222.1) | 3338 / 2688 (1241.9) | 0.89 (0.78; 1.02) |
| **APACHE II score**[2] | | | |
| High (n = 436) | 2874 / 2177 (1320.0) | 4279 / 3633 (1177.9) | 0.87 (0.76; 1.00) |
| Low (n = 476) | 2948 / 2298 (1283.1) | 3835 / 3169 (1210.3) | 0.88 (0.78; 0.99) |
| **Kidney failure** | | | |
| Yes (n = 349) | 1968 / 1637 (1202.5) | 3160 / 2717 (1163.1) | 0.93 (0.81; 1.08) |
| No (n = 563) | 3854 / 2838 (1357.8) | 4954 / 4085 (1212.8) | 0.85 (0.76; 0.95) |
| **Septic shock** | | | |
| Yes (n = 389) | 2815 / 1895 (1485.7) | 4269 / 3350 (1274.3) | 0.88 (0.77; 1.01) |
| No (n = 523) | 3007 / 2580 (1165.3) | 3846 / 3452 (1114.1) | 0.85 (0.75; 0.96) |

Ω Intervention impact tested by the Generalized Linear Model

1 High $SOFA_{24}$ score: scores > 5, Low $SOFA_{24}$ score: scores <=5

2 High APACHE II score: scores >16, Low APACHE II score: scores <=16

n: daily dose of antibacterials, N: number of intensive care unit days of stay *eTable 4. Distribution of antibacterials, including median doses, reported for each period.*

| | Baseline (N= 357) | | Implementation (N = 555) | |
|---|---|---|---|---|
| Antibacterials | n (%) | Median (Q1-Q3) in mg or IU | N (%) | Median (Q1-Q3) in mg or IU |
| Amikacina | 28 (7.8%) | 5 (2-7) | 35 (6.3%) | 3 (2-6) |
| Amoxicilina + acido clavulánico | 5 (1.4%) | 24 (18-36) | 2 (0.4%) | 16 (9-23) |
| Ampicilina | 8 (2.2%) | 49 (40.5-64.975) | 15 (2.7%) | 48 (24-90) |
| Ampicilina + Sulbactam | 146 (40.9%) | 30 (18-41.625) | 236 (42.5%) | 24 (13.125-36) |
| Aztreonam | 4 (1.1%) | 11 (8.8-13.375) | 4 (0.7%) | 26 (21-28.35) |
| Bencilpenicilina | 1 (0.3%) | 1200000* | 1 (0.2%) | 12500000* |
| Bencilpenicilina benzatínica | - | - | 1 (0.2%) | 2400000* |
| Cefazolina | 19 (5.3%) | 15 (9-60) | 14 (2.5%) | 15 (7-27.75) |
| Cefepime | 26 (7.3%) | 18 (10.5-30) | 30 (5.4%) | 18 (12-42) |
| Cefotaxima | 1 (0.3%) | 36* | 4 (0.7%) | 10 (6-18) |
| Ceftazidima | 21 (5.9%) | 15 (6-18) | 20 (3.6%) | 15 (12-27) |
| Ceftazidima + avibactam | 12 (3.4%) | 17.5 (14.75-31.25) | 12 (2.2%) | 31.8 (22.725-40.8) |
| Ceftriaxona | 18 (5.0%) | 8 (4.5-14) | 43 (7.7%) | 12 (4-20) |
| Ciprofloxacina | 11 (3.1%) | 2.5 (1.4-4.4) | 24 (4.3%) | 2.4 (1.5-3.7) |
| Claritromicina | 54 (15.1%) | 4 (2-5) | 66 (11.9%) | 3 (2-5) |
| Clindamicina | 28 (7.8%) | 7.8 (3.3-12.6) | 32 (5.8%) | 7.8 (4.5-11.1) |
| Colistin | 138 (38.7%) | 1.4 (0.8-2.1) | 250 (45.0%) | 1.4 (0.8-2.4) |
| Doxiciclina | 1 (0.3%) | 0.4* | | |
| Fosfomicina (intravenosa) | 3 (0.8%) | 60 (32-86) | 10 (1.8%) | 56 (28-121) |
| Gentamicina | 11 (3.1%) | 1.2 (1-1.4) | 11 (2.0%) | 1 (0.95-1.8) |
| Imipenem | 11 (3.1%) | 9 (6.75-10.25) | 12 (2.2%) | 6 (3.75-10.625) |
| Levofloxacina | 3 (0.8%) | 3.2 (1.85-3.85) | 5 (0.9%) | 3.5 (2.4-4) |
| Linezolid | 15 (4.2%) | 4 (3.6-9.6) | 13 (2.3%) | 6 (3.6-9.6) |
| Meropenem | 110 (30.8%) | 15 (9-24) | 170 (30.6%) | 18 (12-30) |
| Meropenem + vaborbactam | | | 1 (0.2%) | 126* |
| Metronidazol | 12 (3.4%) | 5.25 (4.125-7.5) | 24 (4.3%) | 6 (1.5-12) |
| Nitrofurantoina | 1 (0.3%) | 1.2 (1.2-1.2) | 1 (0.2%) | 1.4* |
| Otros | 36 (10.1%) | 1.65 (0.75-4) | 16 (2.9%) | 2.2 (0.9-7.2) |
| Piperacilina + tazobactam | 120 (33.6%) | 54 (36-82.525) | 224 (40.4%) | 54 (30.875-90) |
| Procaína bencilpenicilina | | | 1 (0.2%) | 24000000* |
| Tigeciclina | 15 (4.2%) | 1 (0.7-1.5) | 39 (7.0%) | 0.7 (0.4-1.4) |
| Trimetoprima + sulfametoxazol | 36 (10.1%) | 5.15 (2.4-10.5) | 19 (3.4%) | 3.2 (1.3-4.5) |
| Vancomicina | 127 (35.6%) | 6 (4-11) | 203 (36.6%) | 7.5 (4-12) |

* Since only one observation was recorded, only the observed value is reported.
IU= international units

*eTable 5. Times from microbiological rescue to antibacterial adjustment and prescription between study periods.*

| Characteristic | Baseline (N = 438) | Intervention (N = 587) | P-value* |
|---|---|---|---|
| | **n/N (%)** | **n/N (%)** | |
| **Requires antibacterial adjustment** | 243/438 (55.5%) | 292/587 (49.7%) | 0.069 |
| **Time ranges from microbiological rescue to antibacterial prescription** | | | <0.001 |
| < 24 h | 111/233 (47.6%) | 181/274 (66.1%) | |
| 24-48 h | 86/233 (36.9%) | 71/274 (25.9%) | |
| >48-72 h | 26/233 (11.2%) | 10/274 (3.6%) | |
| >72hs | 10/233 (4.3%) | 12/274 (4.4%) | |
| Not done | 0/233 (0.0%) | 0/274 (0.0%) | |

*Chi-squared test used

*eTable 6. Antibacterial resistance between study periods.*

| Characteristic | Baseline (N = 438) | Intervention (N = 587) | P-value* |
|---|---|---|---|
| | **n/N (%)** | **n/N (%)** | |
| **Antibacterial resistance** | 158/438 (36.1%) | 210/587 (35.8%) | >0.9 |
| **Antibacterial resistance Type** | | | |
| Methicillin resistance | 18/158 (11.4%) | 47/210 (22.4%) | 0.006 |
| Vancomycin resistance | 10/158 (6.3%) | 6/210 (2.9%) | 0.11 |
| Extended-spectrum beta-lactamase | 33/158 (20.9%) | 38/210 (18.1%) | 0.5 |
| Carbapenemase | 48/158 (30.4%) | 64/210 (30.5%) | >0.9 |
| **Type of carbapenemase** | | | |
| KPC | 31/48 (64.6%) | 33/63 (52.4%) | 0.2 |
| MBL | 22/48 (45.8%) | 31/63 (49.2%) | 0.7 |
| Others | 1/48 (2.1%) | 2/63 (3.2%) | >0.9 |

*Chi- square test used
